# Evaluating the Impact of Financial and Operating Costs on Service Delivery Efficiency in Integrated Care Boards (ICBs) in the UK: A Data Envelopment Analysis and Regression Approach

**DOI:** 10.1101/2025.09.27.25336799

**Authors:** Emmanuel Achori

## Abstract

This research investigates the effects of financial and operating costs and service delivery efficiency within selected Integrated Care Boards (ICBs) in the UK. Utilising a comprehensive dataset, the study employs both regression analysis and Data Envelopment Analysis (DEA) to evaluate the impact of key independent variables of staff cost, efficiency savings, liquidity ratio, interest on lease liabilities, population size, fund, staff turnover, estate investment level, and right-of-use assets on two critical dependent variables (patient waiting time and operating surplus/deficit).

The regression analysis reveals significant effects, highlighting the positive impact of population size and staff turnover on patient waiting times, and the positive correlation between right-of-use assets and operating surplus. Conversely, efficiency savings and increased funding are found to significantly reduce patient waiting times. The DEA identifies variations in efficiency across different ICBs, pinpointing those operating on the efficiency frontier and those with room for improvement.

The findings offer valuable insights for policymakers and healthcare managers aiming to optimise resource allocation, enhance operational efficiency, and ultimately improve patient outcomes within the evolving landscape of integrated care in the UK.

## 1. Introduction

The healthcare landscape globally is under immense pressure, grappling with escalating costs, increasing demand, and the persistent challenge of delivering high-quality, efficient services. In the United Kingdom, the establishment of Integrated Care Boards (ICBs) represents a significant structural reform aimed at fostering collaboration across health and care organisations to improve population health and reduce health inequalities. (NHS England, 2022). These ICBs are tasked with managing substantial budgets and complex operational frameworks, making their financial and operational efficiency paramount to the success of integrated care delivery.

Integrated Care Boards are responsible for planning and commissioning healthcare services for their local populations, bringing together NHS providers, local authorities, and other partners. (Gongora-Salazar, Glogowska, & Greenhalgh, 2022). This integrated approach seeks to move away from fragmented care, promoting a more holistic and person-centered system. However, the effectiveness of ICBs is not solely dependent on their structural design but critically on their ability to manage resources judiciously and optimize operational processes (Kumar & Gangadharan, 2023). Financial sustainability and operational efficiency are intertwined, directly influencing the capacity of ICBs to meet patient needs, particularly in areas such as patient waiting times, which serve as a key indicator of service accessibility and quality.

Patient waiting times have long been a contentious issue within the NHS, reflecting bottlenecks in service provision, resource constraints, and inefficiencies in patient pathways. Prolonged waiting times can lead to poorer health outcomes, reduced patient satisfaction, and increased burden on individuals and their families. Concurrently, the financial health of ICBs, often measured by their operating surplus or deficit, dictates their capacity for investment, innovation, and resilience against unforeseen challenges. A healthy operating surplus allows for strategic reinvestment in services, infrastructure, and workforce development, while persistent deficits can lead to service cuts, staff morale issues, and a downward spiral in care quality.

This research aims to critically evaluate the impact of various financial and operating cost factors on the service delivery efficiency of ICBs. Specifically, we will investigate how independent variables such as staff cost, efficiency savings, liquidity ratio, interest on lease liabilities, population size, fund allocation, staff turnover, estate investment level, and right-of-use assets influence patient waiting time and operating surplus/deficit. By employing a dual methodological approach encompassing regression analysis and Data Envelopment Analysis (DEA), this study seeks to provide a robust and multifaceted understanding of these relationships.

Regression analysis will allow us to quantify the individual impact of each financial and operating variable on patient waiting times and operating surplus/deficit, identifying statistically significant predictors and the direction and magnitude of their influence. This will shed light on which specific cost components or financial indicators are most strongly associated with improvements or deteriorations in service efficiency and financial performance. Furthermore, a scenario analysis will be conducted to explore the hypothetical impact of a significant reduction in staff cost on patient waiting times, offering insights into the potential consequences of such policy interventions.

Complementing the regression analysis, Data Envelopment Analysis (DEA) will be utilised to assess the relative efficiency of individual ICBs. DEA is a non-parametric method that measures the efficiency of multiple decision-making units (DMUs) with multiple inputs and outputs, without requiring a priori assumptions about the functional form of the production frontier. This will enable us to identify best-practice ICBs that operate on the efficiency frontier and benchmark less efficient units, providing actionable insights into areas for operational improvement and resource optimization. By identifying efficient and inefficient ICBs, the study can highlight potential learning opportunities and areas where targeted interventions might yield the greatest benefits.

The findings of this research are expected to contribute significantly to the existing body of knowledge on healthcare management and efficiency. For policymakers, the study will offer evidence-based insights to inform strategic decisions regarding funding models, resource allocation, and policy interventions aimed at improving service delivery. For ICB managers and healthcare professionals, the results will provide a clearer understanding of the financial and operational levers that can be pulled to enhance efficiency, reduce patient waiting times, and ensure financial sustainability. Ultimately, this research seeks to support the overarching goal of integrated care: to deliver more effective, efficient, and equitable healthcare services to the population.

The remainder of this article is structured as follows: Chapter 2 provides a comprehensive literature review, establishing the theoretical and empirical context of the study. Chapter 3 details the methodology, including data sources, variable definitions, and the application of regression analysis and DEA. Chapter 4 presents the results of both the regression and DEA analyses. Chapter 5 discusses these findings, their implications, and the limitations of the study. Finally, Chapter 6 concludes the article with a summary of key insights and recommendations for future research and practice.

## 2. Literature Review

### 2.1. Introduction to Integrated Care Boards (ICBs)

Integrated Care Boards (ICBs) are pivotal NHS organisations in England, established to plan and deliver health and care services for their local populations. (Miller, Glasby, & Dickinson, 2021; Naylor, Cream, Chikwira & Gowar, 2024). They replaced Clinical Commissioning Groups (CCGs) from July 1, 2022, marking a significant shift towards more integrated and collaborative healthcare provision. ICBs are responsible for substantial budgets, often exceeding £100 billion annually, which they allocate to commission services across their geographical areas. Their primary objective is to improve population health, reduce health inequalities, enhance productivity and value for money, and help the NHS support broader social and economic development. This structural reform aims to foster collaboration across health and care organisations, moving away from fragmented care towards a more holistic and person-centered system.

### 2.2. Financial and Operating Costs in ICBs

The financial landscape of ICBs is complex, involving the management of significant public funds. Operating costs, including staff costs, estate investment, and other administrative expenses, are critical components that directly influence an ICB’s ability to deliver efficient services. The efficient management of these costs is crucial for maintaining financial stability and ensuring that resources are optimally utilised for patient care (Naylor, Cream, Chikwira, & Gowar, 2024). Challenges often arise in balancing the need for cost control with the demand for high-quality, accessible services. For instance, some ICBs have faced mandates to reduce running costs, highlighting the constant pressure to achieve efficiency savings. Understanding the dynamics of these costs is essential for assessing the overall financial health and operational capacity of ICBs.

### 2.3. Service Delivery Efficiency

Service delivery efficiency in healthcare refers to the optimal use of resources to achieve desired health outcomes (Sanderson, M., et al., 2023). It encompasses various aspects, including the timely provision of care, effective resource allocation, and the minimisation of waste. In the context of ICBs, efficiency is often measured by metrics such as patient waiting times and the overall financial performance, including operating surplus or deficit (Ozcan, 2014). Factors influencing efficiency can range from staffing levels and their associated costs to the strategic investment in infrastructure and the effective management of assets. Achieving high service delivery efficiency is a core objective for ICBs, as it directly impacts patient satisfaction and health outcomes (Thomson, L. J. M., et al., 2024).

### 2.4. Patient Waiting Time

Patient waiting time is a critical indicator of healthcare service delivery efficiency and patient access. This is measured using 72-weeks patient waiting time in ICB. Prolonged waiting times can lead to adverse health outcomes, reduced patient satisfaction, and increased burden on healthcare systems. (Dorussen, Hansen, Pickering, & Reifler, 2024). Various factors can influence patient waiting times, including staff availability, resource allocation, operational bottlenecks, and the overall demand for services. Understanding the impact of financial and operating costs on patient waiting times is essential for developing strategies to improve access to timely care. This metric serves as a tangible measure of an ICB’s ability to manage its resources effectively to meet patient demand.

### 2.5. Operating Surplus/Deficit

Operating surplus or deficit reflects the financial health and sustainability of an ICB. A surplus indicates that an organisation’s revenues exceed its operating expenses, allowing for reinvestment in services or reserves. Conversely, a deficit suggests that expenses outweigh revenues, potentially leading to financial instability and challenges in service provision. The management of operating costs, including staff costs, efficiency savings, and other financial liabilities, directly impacts an ICB’s ability to achieve a favourable financial position. (Freedman & Wolf, 2023). Analysing the relationship between these costs and the operating surplus/deficit is vital for ensuring the long-term viability of integrated care services and their capacity to deliver consistent care (Smith, Mossialos, Papanicolas & Leatherman, 2009).

### 2.6. Data Envelopment Analysis (DEA) in Healthcare

Data Envelopment Analysis (DEA) is a non-parametric method widely used to measure the relative efficiency of Decision Making Units (DMUs) that consume multiple inputs to produce multiple outputs. In the context of healthcare, DEA has proven to be a valuable tool for evaluating the performance of various entities, including hospitals, clinics, and Integrated Care Boards (ICBs). By constructing a ‘best practice’ frontier based on observed input-output combinations, DEA identifies efficient DMUs and quantifies the inefficiency of others, providing insights into potential areas for improvement. The application of DEA in healthcare efficiency analysis is particularly relevant due to the complex interplay of various resources (inputs) and outcomes (outputs). For instance, inputs can include staff costs, estate investment, and other operational expenditures, while outputs can be patient waiting times (as an inverse measure of efficiency, or transformed), and financial performance like operating surplus. Unlike parametric methods, DEA does not require a pre-specified functional form for the production function, making it flexible for diverse healthcare settings. Several Python libraries are available for implementing DEA, such as <u>Pyfrontier</u>, which facilitate the construction and solution of DEA models, enabling researchers to perform efficiency assessments and identify benchmarks for performance improvement. The use of such tools allows for a robust quantitative analysis of how financial and operating costs influence the efficiency of service delivery in ICBs, ultimately contributing to evidence-based decision-making in healthcare management.

## 3. Methodology

This study employs a quantitative research design to evaluate the impact of financial and operating costs on service delivery efficiency in Integrated Care Boards (ICBs) the UK. The methodology integrates both regression analysis and Data Envelopment Analysis (DEA) to provide a comprehensive understanding of the relationships between the identified variables. The study population consist of forty two (42) ICBs as at 31st August 2025. A sample of five (5) ICBs were selected. The sample ICBs were chosen using the purposive sampling technique. Secondary data was used in this study and data were gathered from the annual reports of the sampled ICBs for the period 2022 to 2024-2025.

### 3.2. Variables

#### Dependent Variables

i. **Patient Waiting Time (PWT):** This variable represents the average waiting time for patients within each ICB. It serves as a key indicator of service delivery efficiency, with lower values indicating better performance. For DEA, this variable was transformed into its inverse (1/PWT) to align with the output maximisation principle, where higher values denote better outcomes.
ii. **Operating Surplus/Deficit (OSD):** This financial metric reflects the difference between an ICB’s operating revenues and operating expenses. A positive value indicates a surplus, while a negative value indicates a deficit. It is a crucial measure of financial health and sustainability. For DEA, negative values were adjusted by adding a constant to ensure all values were positive, as required by the model.

#### Independent Variables

i. **Staff Cost (SC):** Represents the total expenditure on staff within an ICB. This is a major operating cost and a significant input in healthcare service delivery.
ii. **Efficiency Savings (ES):** This variable quantifies the savings achieved through efficiency initiatives. It is expected to have a positive impact on efficiency and financial performance.
iii. **Liquidity Ratio (LR):** A financial ratio indicating an ICB’s ability to meet its short-term obligations. It reflects financial stability.
iv. **Interest on Lease Liabilities (ILL):** The cost incurred on leased assets, representing a financial operating cost.
v. **Population Size (PS):** The size of the population served by the ICB. This variable accounts for the demand-side pressure on service delivery.
vi. **Fund (FD):** Represents the total funding allocated to the ICB, reflecting the available financial resources.
vii. **Staff Turnover (ST):** The rate at which staff leave and are replaced within an ICB. High turnover can impact operational stability and efficiency.
viii. **Estate Investment Level (EIL):** The level of investment in physical infrastructure and properties. This can influence service capacity and quality.
ix. **Right-of-use Assets (UoA):** Assets recognised on the balance sheet for which the ICB has the right to use for a period, typically under a lease agreement. This reflects the asset base available for operations.

All variables were transformed to using natural logarithms.

### 3.3. Analytical Methods

#### 3.3.1. Regression Analysis

Ordinary Least Squares (OLS) regression was employed to analyse the linear impact between the independent variables and each of the dependent variables (Patient Waiting Time and Operating Surplus/Deficit). Two separate regression models were constructed:

- **Model 1: Patient Waiting Time (PWT) as Dependent Variable:** This model aimed to identify which financial and operating cost factors significantly influence patient waiting times. The independent variables included staff cost, efficiency savings, liquidity ratio, interest on lease liabilities, population size, fund, staff turnover, estate investment level, and right-of-use assets.
- **Model 2: Operating Surplus/Deficit (OSD) as Dependent Variable:** This model investigated the impact of the same set of independent variables on the financial performance (operating surplus/deficit) of ICBs.

For both models, statistical significance was assessed using p-values, and the overall explanatory power was evaluated using R-squared and Adjusted R-squared values. The presence of multi-collinearity was also noted and considered in the interpretation of results.

#### 3.3.2. Data Envelopment Analysis (DEA)

Data Envelopment Analysis (DEA) was utilised to measure the relative efficiency of the ICBs. An input-oriented CCR (Charnes, Cooper, Rhodes, 1978) model was selected, assuming constant returns to scale. This model aims to minimise inputs while maintaining or exceeding current output levels. The ICBs were treated as Decision Making Units (DMUs).

- **Inputs for DEA:**
  - Staff Cost
  - Interest on Lease Liabilities
  - Estate Investment Level
  - Right-of-use Assets
- **Outputs for DEA:**
  - Inverse of Patient Waiting Time (1/PATIENT_WAITING_TIME)
  - Operating Surplus/Deficit (transformed to ensure positivity)
  - Efficiency Savings
  - Fund

The <u>Pyfrontier</u> Python library was used to implement the DEA model. The analysis generated efficiency scores for each DMU, with a score of 1.0 indicating full efficiency relative to the other DMUs in the sample. Scores greater than 1.0 indicate inefficiency, suggesting potential for improvement in resource utilisation.

### 3.4. Scenario Analysis

A scenario analysis was conducted to explore the hypothetical impact of a 50% reduction in staff cost on patient waiting time. This involved adjusting the <u>staff_cost</u> variable in the dataset by 50% and then using the coefficients from the Patient Waiting Time regression model to predict the new average patient waiting time. This analysis aimed to provide insights into the potential consequences of significant cost-cutting measures, while also highlighting the limitations of linear models for extreme policy changes.

## 4. Results

### 4.1. Impact of Independent Variables on Patient Waiting Time (Regression Analysis)

The Ordinary Least Squares (OLS) regression analysis was conducted to evaluate the impact of various financial and operating costs on patient waiting time. The model achieved a high R-squared value of 0.922, indicating that approximately 92.2% of the variance in patient waiting time can be explained by the independent variables included in the model. The adjusted R-squared of 0.781 further supports the model’s strong explanatory power, even after accounting for the number of predictors. The overall model is statistically significant, as indicated by the F-statistic of 6.561 and a p-value of 0.0260 (p < 0.05).

Here’s a detailed breakdown of the impact of each independent variable on patient waiting time:

- **Staff Cost (coef: 2.8452, p-value: 0.258)**: The coefficient for staff cost is positive, suggesting that an increase in staff cost is associated with an increase in patient waiting time. However, with a p-value of 0.258, this relationship is not statistically significant at conventional levels (e.g., 0.05 or 0.10). This might imply that while there’s a positive trend, other factors or a larger dataset might be needed to confirm a significant direct impact.
- **Efficiency Savings (coef: −14.8781, p-value: 0.035)**: This variable shows a statistically significant negative impact on patient waiting time (p < 0.05). A negative coefficient indicates that higher efficiency savings are associated with a reduction in patient waiting time. This is an expected and desirable outcome, as efficiency improvements should ideally lead to smoother operations and shorter waiting periods for patients.
- **Interest on Lease Liabilities (coef: −0.4819, p-value: 0.362)**: The coefficient is negative, suggesting that higher interest on lease liabilities might be associated with lower patient waiting times. However, this relationship is not statistically significant (p = 0.362).
- **Population Size (coef: 35.1018, p-value: 0.012)**: Population size has a highly significant positive impact on patient waiting time (p < 0.05). A large positive coefficient indicates that an increase in the population served by an ICB is strongly associated with a substantial increase in patient waiting time. This highlights the pressure that larger populations place on healthcare service delivery.
- **Estate Investment Level (coef: 0.4546, p-value: 0.122)**: The positive coefficient suggests that higher investment in estates is associated with increased patient waiting times. While not statistically significant at the 0.05 level, it is close to significance at the 0.10 level (p = 0.122). This counter-intuitive result might warrant further investigation, as one might expect estate investments to improve service capacity and reduce waiting times. It could indicate that investments are not yet yielding their full benefits, or are directed towards areas not directly impacting waiting times.
- **Staff Turnover (coef: 2.3280, p-value: 0.058)**: Staff turnover shows a positive impact on patient waiting time, meaning higher turnover is associated with longer waiting times. This relationship is statistically significant at the 0.10 level (p = 0.058), suggesting that frequent changes in staff can disrupt service continuity and efficiency, leading to increased waiting times.
- **Liquidity Ratio (coef: 0.3822, p-value: 0.648)**: The liquidity ratio has a positive coefficient, but its impact on patient waiting time is not statistically significant (p = 0.648).
- **FUND (coef: −12.8948, p-value: 0.021)**: This variable has a statistically significant negative impact on patient waiting time (p < 0.05). A negative coefficient suggests that higher funding is associated with a reduction in patient waiting time, which is an expected and positive outcome.
- **Right-of-use Assets (coef: 1.0522, p-value: 0.154)**: The positive coefficient suggests that an increase in right-of-use assets is associated with increased patient waiting times, though this relationship is not statistically significant (p = 0.154).

### 4.2. Impact of Independent Variables on Operating Surplus/Deficit (Regression Analysis)

The second OLS regression model was developed to understand the drivers of operating surplus or deficit in ICBs. This model also demonstrates strong explanatory power, with an R-squared of 0.917 and an adjusted R-squared of 0.766. The F-statistic of 6.099 and a corresponding p-value of 0.0303 (p < 0.05) confirm the overall significance of the model.

Here is a detailed analysis of each independent variable’s impact on the operating surplus/deficit:

- **Staff Cost (coef: −6.0488, p-value: 0.337)**: The negative coefficient suggests that higher staff costs are associated with a lower operating surplus (or a larger deficit). However, this relationship is not statistically significant (p = 0.337).
- **Efficiency Savings (coef: −24.2817, p-value: 0.126)**: The coefficient is negative, indicating that higher efficiency savings are linked to a lower operating surplus. This is a counter-intuitive finding and is not statistically significant at the 0.05 level, although it is approaching significance at the 0.10 level (p = 0.126). This could suggest that the costs of achieving efficiency savings are not being fully offset by the benefits within the same period, or that savings are being reinvested elsewhere.
- **Interest on Lease Liabilities (coef: 1.5380, p-value: 0.265)**: The positive coefficient suggests a positive relationship with the operating surplus, but this is not statistically significant (p = 0.265).
- **Population Size (coef: 13.3303, p-value: 0.589)**: The coefficient is positive, but the relationship is not statistically significant (p = 0.589).
- **Estate Investment Level (coef: −0.5266, p-value: 0.437)**: The negative coefficient suggests that higher estate investment is associated with a lower operating surplus, but this is not statistically significant (p = 0.437).
- **Staff Turnover (coef: 3.7344, p-value: 0.184)**: The positive coefficient indicates that higher staff turnover is associated with a higher operating surplus. This is another counter-intuitive finding, though not statistically significant (p = 0.184). It could be that lower-cost staff are replacing more expensive staff, leading to short-term cost reductions.
- **Liquidity Ratio (coef: −1.7282, p-value: 0.429)**: The negative coefficient suggests a negative relationship with the operating surplus, but this is not statistically significant (p = 0.429).
- **FUND (coef: 14.4862, p-value: 0.201)**: The positive coefficient indicates that higher funding is associated with a higher operating surplus, but this relationship is not statistically significant (p = 0.201).
- **Right-of-use Assets (coef: 5.1445, p-value: 0.024)**: This variable has a statistically significant positive impact on the operating surplus (p < 0.05). A higher value of right-of-use assets is strongly associated with a higher operating surplus. This could be due to the nature of how these assets are financed and accounted for, potentially reflecting a more favourable financial position.

### 4.3. Data Envelopment Analysis (DEA) Results

The Data Envelopment Analysis (DEA) was performed using an input-oriented CCR (Charnes, Cooper, Rhodes, 1978) model to assess the relative efficiency of the Integrated Care Boards (ICBs) over the specified years. The CCR model assumes constant returns to scale, meaning that an increase in inputs leads to a proportional increase in outputs.

The efficiency scores range from 1.0000 to approximately 1.0477. A score of 1.0000 indicates that the Decision Making Unit (DMU) is operating on the efficiency frontier, meaning it is fully efficient relative to the other DMUs in the sample. DMUs with scores greater than 1.0000 are considered inefficient, indicating that they could achieve the same level of outputs with fewer inputs, or produce more outputs with their current inputs, if they were to operate as efficiently as the frontier DMUs.

#### Key Observations from DEA Scores

- **Efficient ICBs (Score = 1.0000):** Most ICBs, including NHS Kent & Medway, NHS Derby and Derbyshire, North Central London, and Humber & North Yorkshire, demonstrate an efficiency score of 1.0000 across all years (2022-23, 2023-24, 2024-25). This suggests that these ICBs are operating at their optimal capacity given their inputs and outputs, serving as benchmarks for the less efficient units.
- **Inefficient ICBs (Score > 1.0000):** North East London shows efficiency scores greater than 1.0000 for all three years:
  - North East London_2024-25: 1.0398
  - North East London_2023-24: 1.0440
  - North East London_2022-23: 1.0477
- These scores indicate that North East London ICB is relatively inefficient compared to the other ICBs in the sample. For instance, a score of 1.0398 for 2024-25 suggests that this ICB could potentially reduce its inputs by approximately 3.98% (1 - 1/1.0398) while maintaining its current output levels, or increase its outputs by 3.98% with its current inputs, to reach the efficiency frontier. The consistent inefficiency across years for North East London suggests systemic issues or areas for improvement in resource utilisation or service delivery processes.

#### Implications

The DEA results provide valuable insights into the relative performance of ICBs. Efficient ICBs can serve as models for best practices, while inefficient ICBs can identify areas where they need to improve. Further investigation into the operational differences between efficient and inefficient ICBs could reveal specific strategies for enhancing service delivery efficiency and financial management.

## 5. Discussion

This study aimed to evaluate the impact of financial and operating costs on service delivery efficiency in Integrated Care Boards (ICBs) in the UK, using patient waiting time and operating surplus/deficit as key performance indicators. Our findings, derived from both regression analysis and Data Envelopment Analysis (DEA), offer valuable insights into the complex interplay of these factors within the integrated care landscape.

### 5.1. Key Findings from Regression Analysis

The regression analysis provided a quantitative understanding of how individual financial and operating variables influence patient waiting times and operating surplus/deficit. For patient waiting times, the most significant findings were:

- **Efficiency Savings:** The statistically significant negative coefficient for efficiency savings on patient waiting time is a crucial finding. It suggests that efforts to improve efficiency directly translate into shorter waiting periods for patients. This aligns with the core objectives of ICBs to optimize resource utilization and streamline service delivery. It underscores the importance of robust efficiency programs and their potential to yield tangible benefits for patients.
- **Population Size:** The strong positive impact of population size on patient waiting time highlights a fundamental challenge for ICBs. As the population served grows, the demand for healthcare services inevitably increases, placing greater pressure on existing resources and infrastructure. This finding suggests that ICBs serving larger populations may require proportionally greater resources or more innovative service delivery models to maintain acceptable waiting times. It also points to the need for proactive planning and investment in areas with demographic growth.
- **Staff Turnover:** The positive and near-significant impact of staff turnover on patient waiting time is a critical operational concern. High staff turnover can disrupt team cohesion, lead to a loss of institutional knowledge, increase recruitment and training costs, and ultimately impair service continuity and efficiency. This finding emphasises the importance of workforce stability and effective human resource management strategies within ICBs to mitigate the adverse effects on patient access.
- **Funding:** The significant negative impact of overall funding on patient waiting time is an expected yet important validation. Adequate funding provides ICBs with the necessary resources to expand capacity, invest in new technologies, and recruit and retain staff, all of which can contribute to reducing waiting times. This reinforces the argument for appropriate and sustainable funding models for integrated care.

For operating surplus/deficit, the most notable finding was:

- **Right-of-use Assets:** The statistically significant positive impact of right-of-use assets on operating surplus is an interesting financial insight. This could reflect the accounting treatment of these assets under IFRS 16, where the recognition of a right-of-use asset and a corresponding lease liability on the balance sheet can impact financial ratios and perceived financial health. It might also indicate that ICBs effectively leverage leased assets to generate revenue or reduce other operational costs, thereby contributing positively to their financial bottom line. Further investigation into the specific nature and utilization of these assets would be beneficial.

### 5.2. Insights from Data Envelopment Analysis (DEA)

The DEA provided a complementary perspective by assessing the relative efficiency of individual ICBs. The identification of a clear efficiency frontier, with several ICBs consistently achieving a score of 1.0000, offers valuable benchmarks. These efficient ICBs demonstrate best practices in converting inputs into outputs, suggesting that their operational models, resource allocation strategies, and management practices are highly effective relative to their peers.

Conversely, the consistent inefficiency observed in North East London ICB across all years (scores > 1.0000) highlights a significant opportunity for improvement. The inefficiency scores quantify the extent to which this ICB could potentially reduce its inputs or increase its outputs to reach the efficiency frontier. For example, the 2024-25 score of 1.0398 implies a potential for nearly 4% improvement in input utilization or output generation. This finding warrants a deeper, qualitative investigation into the operational and strategic differences between North East London and the efficient ICBs. Such an investigation could uncover specific inefficiencies in resource deployment, service delivery processes, or strategic planning that contribute to its lower relative efficiency.

The DEA results underscore the importance of peer learning and benchmarking within the ICB system. By studying the practices of efficient ICBs, less efficient units can identify actionable strategies for improvement. This could involve adopting similar workforce management practices, optimizing procurement processes, or re-evaluating investment strategies to enhance overall productivity.

## 6. Conclusion

This study has provided a comprehensive evaluation of the impact of financial and operating costs on service delivery efficiency in Integrated Care Boards (ICBs) in the UK, utilizing a dual methodological approach of regression analysis and Data Envelopment Analysis (DEA). Our findings illuminate several critical factors influencing patient waiting times and operating surplus/deficit, offering valuable insights for enhancing the effectiveness and sustainability of integrated care.

The regression analysis revealed that efficiency savings (ES) and increased funding (FD) are significant drivers in reducing patient waiting times, underscoring the importance of strategic financial management and adequate resource allocation. Conversely, population size (PS) and staff turnover (ST) were found to significantly increase patient waiting times, highlighting the inherent challenges of managing demand in growing populations and the critical need for workforce stability. For financial performance, right-of-use assets (UoA) emerged as a significant positive predictor of operating surplus (OSD), suggesting effective asset utilization or favourable accounting treatments.

The Data Envelopment Analysis (DEA) complemented these findings by identifying a clear efficiency frontier among the ICBs. Several ICBs consistently demonstrated full efficiency, serving as benchmarks for best practices in resource conversion. The consistent inefficiency observed in North East London ICB across the years, points to specific areas where operational improvements and strategic re-evaluation are needed to optimize resource utilization and service delivery. This highlights the potential for peer learning and targeted interventions to improve overall system efficiency.

In conclusion, achieving optimal service delivery efficiency and financial sustainability in ICBs requires a balanced approach that prioritizes strategic efficiency initiatives, secures adequate funding, addresses the pressures of population growth, and fosters a stable and supported workforce. Future research should aim to expand the dataset, incorporate qualitative factors, and explore more sophisticated modeling techniques to further refine our understanding of these critical relationships. By leveraging these insights, policymakers and healthcare managers can make more informed decisions to strengthen integrated care and improve health outcomes for all.

### 5.5. Policy Implications and Recommendations

The findings of this study carry significant policy implications for the UK healthcare system, particularly for the strategic direction and operational management of ICBs. The identified relationships between financial and operating costs and service delivery efficiency suggest several areas where targeted policy interventions could yield substantial benefits.

Firstly, the strong negative correlation between efficiency savings and patient waiting time underscores the critical importance of sustained and well-executed efficiency programs. Policymakers should continue to incentivise and support ICBs in identifying and implementing efficiency improvements. However, the counter-intuitive finding that efficiency savings were negatively associated with operating surplus (though not statistically significant) warrants further investigation. This could imply that current efficiency metrics or accounting practices might not fully capture the long-term financial benefits, or that the costs associated with implementing efficiency initiatives temporarily outweigh immediate financial gains. Future policy should aim for a more holistic evaluation of efficiency programs, considering both operational and financial outcomes over a longer timeframe.

Secondly, the significant positive impact of population size on patient waiting times highlights the need for a differentiated approach to resource allocation across ICBs. A ‘one-size-fits-all’ funding model may not adequately address the disproportionate pressures faced by ICBs serving larger or rapidly growing populations. Policy should consider mechanisms for equitable resource distribution that account for demographic factors, potentially through weighted capitation formulas that better reflect the demand-side pressures. Furthermore, ICBs in high-growth areas may require additional support for infrastructure development and workforce planning to proactively manage increasing patient demand.

Thirdly, the impact of staff turnover on patient waiting times is a critical workforce policy concern. High turnover rates not only disrupt service continuity but also incur significant costs related to recruitment, training, and temporary staffing. Policies aimed at improving staff retention, such as enhanced professional development opportunities, improved working conditions, competitive remuneration, and robust wellbeing support, are essential. Investing in workforce stability can lead to more experienced teams, better patient flow, and ultimately, reduced waiting times. This also aligns with broader NHS workforce strategies aimed at creating a more sustainable and resilient healthcare workforce.

Fourthly, the positive relationship between funding and reduced patient waiting times reinforces the argument for adequate and stable investment in the NHS. While efficiency is paramount, it cannot compensate for chronic underfunding. Policymakers must ensure that ICBs receive sufficient financial allocations to meet the healthcare needs of their populations, enabling them to invest in capacity expansion, technological advancements, and innovative models of care that directly address waiting list backlogs. The study suggests that increased funding, when coupled with effective management, can be a powerful lever for improving patient access.

Finally, the DEA results provide a powerful tool for benchmarking and peer learning. Policy should facilitate platforms and processes for efficient ICBs to share their best practices with less efficient counterparts. This could involve structured mentorship programs, knowledge exchange networks, and performance improvement collaboratives. By understanding the operational and strategic choices of frontier ICBs, those with lower efficiency scores can develop targeted improvement plans. Regulatory bodies could also use DEA results to identify ICBs requiring specific support or intervention to enhance their efficiency and resource utilization.

In summary, policy recommendations stemming from this research advocate for a multi-pronged approach: fostering genuine efficiency improvements, adapting resource allocation to population needs, prioritizing workforce stability, ensuring adequate funding, and leveraging benchmarking for continuous improvement. These measures, if implemented thoughtfully, can contribute to a more resilient, efficient, and patient-centered integrated care system in the UK.

## Data Availability

Secondary data - Annual Financial Statements of the selected HNS ICBs

